# Altered IgG4 Antibody Response to Repeated mRNA versus Protein COVID Vaccines

**DOI:** 10.1101/2024.01.17.24301374

**Authors:** Raj Kalkeri, Mingzhu Zhu, Shane Cloney-Clark, Joyce S. Plested, Anand Parekh, Drew Gorinson, Rongman Cai, Soham Mahato, Pradhipa Ramanathan, L. Carissa Aurelia, Kevin John Selva, Anthony M. Marchese, Louis Fries, Amy W. Chung, Lisa M. Dunkle

## Abstract

Repeated mRNA SARS-CoV-2 vaccination has been associated with increases in the proportion of IgG4 in spike-specific antibody responses and concurrent reductions in Fcγ-mediated effector functions that may limit control of viral infection. Here, we assessed anti-Spike total IgG, IgG1, IgG2, IgG3 and IgG4, and surrogate markers for antibody-dependent cellular phagocytosis (ADCP, FcγRIIa binding), antibody-dependent cellular cytotoxicity (ADCC, FcγRIIIa binding), and antibody-dependent complement deposition (ADCD, C1q binding) associated with repeated SARS-CoV-2 vaccination with NVX-CoV2373 (Novavax Inc., Gaithersburg, MD). The NVX-CoV2373 protein vaccine did not induce notable increases in spike-specific IgG4 or negatively impact surrogates for Fcγ effector responses. Conversely, repeated NVX-CoV2373 vaccination uniquely enhanced IgG3 responses which are known to exhibit strong affinity for FcγRIIIa and have previously been linked to potent neutralization of SARS-CoV-2. Subsequent investigations will help to understand the immunological diversity generated by different SARS-CoV-2 vaccine types and have the potential to reshape public health strategies.

## Introduction

The four unique subclasses of human IgG, numbered in order of relative abundance, include IgG1, IgG2, IgG3, and IgG4^1,2^. IgG1 and IgG3 are the major contributors to rapid IgG responses to protein and membrane antigens, and are active in viral neutralization, although some differences in IgG subclass responses have been identified in the context of SARS-CoV-2 infection and vaccination. Indeed, IgG3 response to SARS-CoV-2 infection has been highlighted by an analysis of convalescent plasma, showing that while IgG3 accounted for 12% of total anti-Spike (anti-S) protein IgG, it contributed to approximately 80% of total live SARS-CoV-2 neutralizing activity^3^. In addition, IgG1, and especially IgG3, bind FcγRIIa, FcγRIIIa and C1q, thereby supporting antibody-dependent cellular phagocytosis (ADCP), antibody-dependent cytotoxicity (ADCC), and antibody-dependent complement deposition (ADCD)^2^.

Repeated mRNA SARS-CoV-2 vaccination has been associated with significant increases in the proportion of immunoglobulin G4 (IgG4) in spike-specific responses and reductions in Fc-mediated ADCP and ADCD that may limit control of viral infection^4-8^. After repeated mRNA SARS-CoV-2 vaccination, IgG3 was observed to reach peak titers after a second dose, and steadily declined thereafter to significantly lower levels after the third and fourth dose vaccinations, while IgG4 increased^5^. IgG4 is generally in low abundance (0–5% of total IgG) but may slowly increase over time due to repeated or excessive exposure to some antigens^1,2,8^. Although repeated antigen exposure seems necessary, it is insufficient to induce IgG4, as prolonged activation of IL-10–expressing CD4^+^ T-cells or expression of other anti-inflammatory cytokines is also necessary^1^. Increased concentrations of IgG4 have been associated with immunosuppression and poor clinical outcomes of COVID-19, and while generally regarded as anti-inflammatory, may contribute to some autoimmune disorders and inflammatory IgG4-related diseases^1,8^. Following repeated mRNA vaccination, IgG4 was observed to increase from 0.04% of total SARS-CoV-2 spike–specific IgG after two doses to 19.27% after three doses^4^.

## Methods

We assessed IgG subclass and Fc effector function profiles following repeated vaccination with the recombinant Spike (rS) protein SARS-CoV-2 vaccine (NVX-CoV2373, Novavax, Inc.) in comparison to a single rS vaccination following repeated mRNA vaccinations. Serum concentrations of anti–ancestral (Wuhan) rS–specific total IgG, IgG1, IgG2, IgG3, and IgG4, and surrogate ADCP (FcγRIIa binding), surrogate ADCC (FcγRIIIa binding), and ADCD (C1q binding) were measured. Participant sera from studies 2019nCoV-307 (ClinicalTrials.gov: NCT05463068) and 2019nCoV-301 (ClinicalTrials.gov: NCT04611802) were included in the analysis; two groups from 2019nCoV-307 who received three homologous doses of Moderna (mRNA-1273, *n* = 10) or Pfizer (BNT162b2, *n* = 10) mRNA vaccine followed by one dose of NVX-CoV2373, and a third group from 2019nCoV-301 who received four homologous doses of Novavax (*n* = 18). Serum was collected ≥6 months after the last dose for homologous three-dose series and ∼4 weeks after the fourth-dose for the four-dose series.

Anti-S proteins IgG1, IgG2, IgG3, and IgG4, and total anti-S IgG were measured for each serum sample by IgG subclass quantitative enzyme-linked immunosorbent assay (ELISA). SARS-CoV-2 rS proteins (produced at Novavax, Inc., Gaithersburg, MD, USA) were used to coat ELISA plates. The reference standards for ELISA were as follows; SARS-CoV-2 Spike RBD human IgG monoclonal antibody (Acro Biosystems, Cat# RAS009-C02) for total IgG, anti-SARS-CoV-2 Spike RBD human IgG1 monoclonal antibody (Acro Biosystems, Cat# SPD-M265) for IgG1, anti-SARS-CoV-2 spike RBD human IgG2 monoclonal antibody (Acro Biosystems, Cat# SPD-M400a), anti-SARS-CoV-2 spike RBD human IgG3 monoclonal antibody (Acro Biosystems, Cat# SPD-M401a), and anti-SARS-CoV-2 Spike RBD human IgG4 monoclonal antibody (Acro Biosystems, Cat# SPD-M402a) for IgG4. To determine ADCP, ADCC, and ADCD, surrogate SARS-CoV-2–specific antibody Fc functional assays were performed as previously described 9. The surrogate Fc multiplex assays have been shown to correlate strongly with cell-based ADCP and ADCC functional assays^9^. Briefly, SARS-Cov-2 spike trimer (SinoBiological Cat# 40589) coupled multiplex beads, were used to profile antigen-specific FcγR activation or C1q (complement) deposition as a high-throughput surrogate to assess ADCP by soluble FcγRIIa dimer binding, ADCC by soluble FcγRIIIa dimer binding (both expressed in house at University of Melbourne) or ADCD by C1q binding (MP Biomedicals, Irvine, CA, USA).

## Results

Total anti-S IgG and IgG1 levels following three homologous doses of mRNA or NVX-CoV2373 were similar, although NVX-CoV2373 induced somewhat higher levels, and the fourth dose of NVX-CoV2373 led to increased responses in each group (Fig. 1A). Compared with recipients of prior mRNA vaccine, anti-S IgG3 levels were markedly higher (>10-fold) after three or four homologous doses of NVX-CoV2373. By contrast, much higher anti-S IgG4 levels (>75-fold) were observed following repeated mRNA vaccination, but not after three or four homologous doses of NVX-CoV2373 (Fig. 1A). The fourth dose of NVX-CoV2373 also appeared to enhance surrogate signals for ADCP, ADCC, and ADCD activities in recipients of prior mRNA vaccine, though the effect was greater after a fourth homologous dose of NVX-CoV2373 (Fig. 1B).

**Figure 1.**
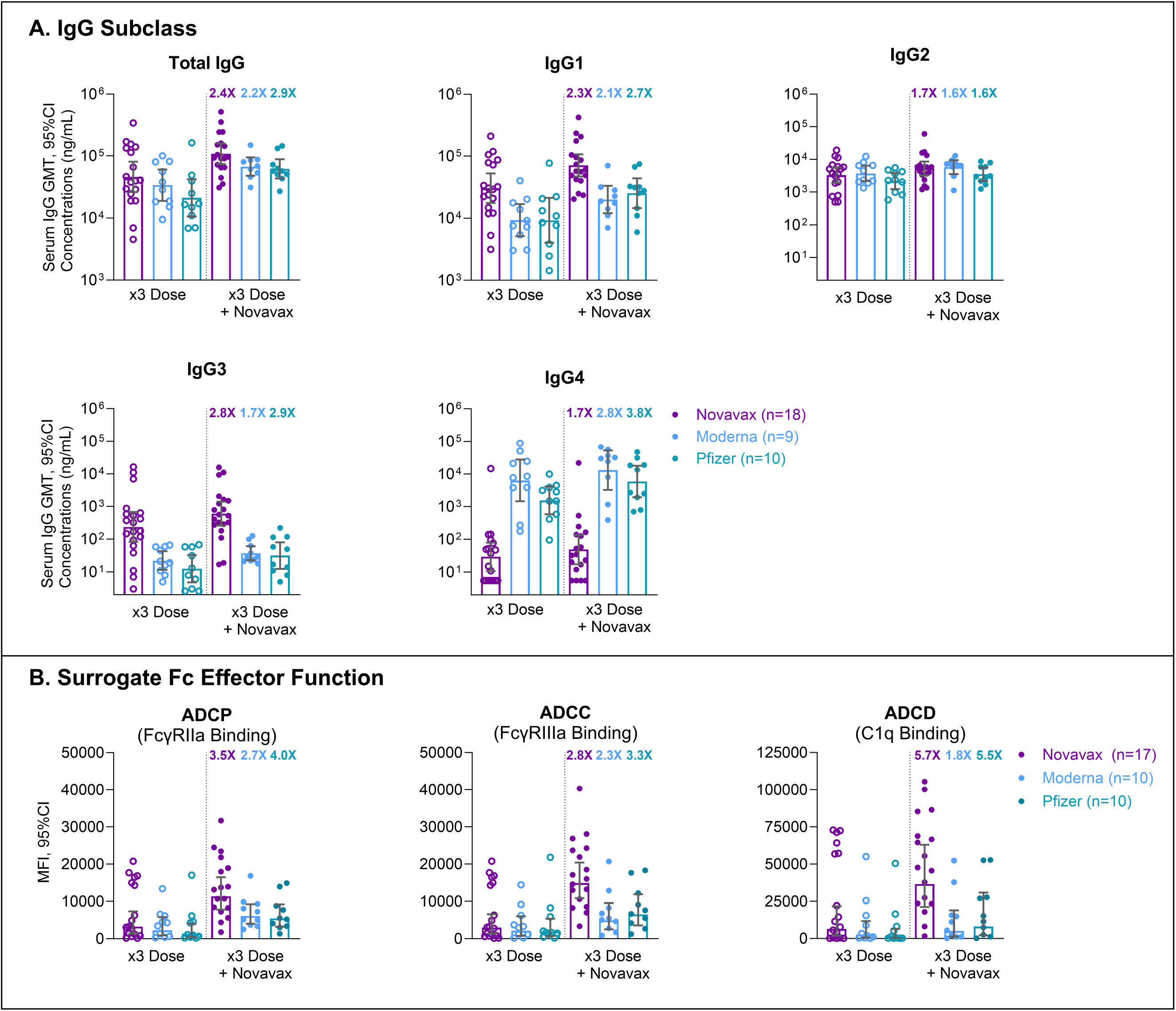
IgG Subclass Serum Concentrations and Fc Effector Function Responses After Repeated COVID Vaccination. Randomly selected serum samples from COVID vaccine recipients included participants from 2019nCoV-307 (ClinicalTrials.gov: NCT05463068) who received three homologous doses of Moderna (mRNA-1273, blue, *n* = 10) or Pfizer (BNT162b2, teal, *n* = 10) mRNA vaccine followed by one dose of Novavax (NVX-CoV2373), and from 2019nCoV-301 (ClinicalTrials.gov: NCT04611802) who received four homologous doses of Novavax (purple, *n* = 18). Serum was collected ≥6 months after the last dose for homologous three-dose samples (open circle) and ∼4 weeks after fourth dose samples (filled circle). For each vaccine type, the GMFR between the third and fourth doses is indicated in bolded text above the fourth-dose data points. **(A)** Serum concentrations of anti–ancestral (Wuhan) rS–specific total IgG, IgG1, and IgG4 were measured using quantitative ELISA and reported as GMT with 95% CI. One subject (Moderna) was excluded due to a failed logic check. **(B)** ADCP, ADCC, and ADCD were analyzed using previously described surrogate SARS-CoV-2 ancestral rS–specific antibody Fc functional multiplex assays assessing median fluorescence intensity of FcγRIIa, FcγRIIIa, and C1q binding, respectively. One subject (Novavax) was excluded due to missing data (x3 Dose + Novavax). SAS^®^ 9.4 software (SAS Institute Inc., Cary, NC), PROC SGPLOT was used to detect far-outliers based on the log-transformed assay values, defined as Far-outliers = Observations > Q3 + 3*IQR or < Q1 – 3*IQR; none were found or excluded. Abbreviations: ADCC, antibody-dependent cellular cytotoxicity; ADCD, antibody-dependent complement deposition; ADCP, antibody-dependent cellular phagocytosis; CI, confidence interval; C1q complement component 1q; ELISA, enzyme-linked immunosorbent assay; FcγRIIa, Fcγ Receptor IIa; FcγRIIIa, Fcγ Receptor IIIa; GMFR, geometric mean fold rise; GMT, geometric mean titer; IgG, Immunoglobulin G; MFI, median fluorescent intensity; rS, recombinant spike protein; 3x, three-dose.

## Discussion

The clinical importance of SARS-CoV-2–specific Fc-mediated responses (i.e., ADCP, ADCD, and ADCC), which are poorly engaged by IgG4, is rapidly gaining appreciation^4,6-8,10^. Fcγ-dependent effector functions can provide additional mechanisms for virus control to complement neutralization, and these may be important for the promotion of vaccine-mediated cross-protection to evolving SARS-CoV-2 variants^10^. Here, we report that the NVX-CoV2373 protein vaccine does not appear to induce notable increases in IgG4, even after multiple exposures, or impair Fcγ-dependent effector responses as observed with mRNA vaccines. Instead, NVX-CoV2373 drove proportional increases in IgG3, perhaps the most potent SARS-CoV-2 neutralizing antibody subclass^3^, and enhanced surrogate ADCP, ADCD, and ADCC activity. The impact of additional doses with updated mRNA and protein-based XBB.1.5 formulation vaccines is currently under study and represents a potentially important area for future COVID-19 vaccine research. Ongoing investigations of these effects on IgG subclasses and cellular functions will help to elucidate the immunological diversity generated by different SARS-CoV-2 vaccine platforms.

## Data Availability

The datasets generated during and/or analyzed for this publication are not available at this time.

## Acknowledgements

The Sponsor acknowledges the contributions and participation of all 2019nCoV-301 and 2019nCoV-307 study volunteers, principal investigators, and investigative site personnel who contributed to the success of the studies. We thank Bruce D. Wines and P. Mark Hogarth, Burnet Institute, Melbourne, for sharing their Fcγ Receptor dimer plasmids. The authors would also like to thank Hadi Beyhaghi and Muruga Vadivale for scientific contributions to the development of the letter, and Seth Toback, Matt Rousculp, and Brandy Warren for cross-functional assistance in project coordination.

## Author Contributions

Conceptualization: R.K., M.Z., A.W.C., S.C., J.S.P., L.M.D., and L.F. Investigation: R.K., M.Z., A.W.C., S.C., J.S.P., A.P., D.G., R.C., S.M., P.R., L.C.A., and K.J.S. Data Analysis: R.K., M.Z., A.W.C., S.C., A.P., D.G., R.C., S.M. P.R., A.M.M., and J.S.P. Visualization: R.K. A.W.C., A.M.M., L.F., and L.M.D.

Writing—original draft: A.M.M. Writing—review and editing: All authors. Project Administration: R.K.

## Author Disclosures

R.K., M.Z., S.C-C, J.S.P., A.P., D.G., R.C., S.M., A.M.M., L.F., and L.M.D. are the employees and stockholders of Novavax, Inc.

P.R., L.C.A., K.J.S., and A.W.C. are the employees of the Department of Microbiology and Immunology, University of Melbourne, at the Peter Doherty Institute for Infection and Immunity. A.W.C. received grant funding from NHMRC, MRFF, and NIH. PR., L.C.A., and K.J.S. declare no conflicts/disclosure information.

### Additional Funding Information

This study was funded by Novavax, Inc.

Samples from 2019nCoV-301 (ClinicalTrials.gov: NCT04611802) were included in the present study. 2019nCoV-301 was funded by the Office of the Assistant Secretary for Preparedness and Response, Biomedical Advanced Research and Development Authority (BARDA; contract Operation Warp Speed: Novavax Project Agreement number 1 under Medical CBRN [Chemical, Biological, Radiological, and Nuclear] Defense Consortium base agreement no. 2020-530; Department of Defense no.

W911QY20C0077); and the National Institute of Allergy and Infectious Diseases (NIAID), National Institutes of Health. The NIAID provides grant funding to the HIV Vaccine Trials Network (HVTN) Leadership and Operations Center (UM1 AI68614), the HVTN Statistics and Data Management Center (UM1 AI68635), the HVTN Laboratory Center (UM1 AI68618), the HIV Prevention Trials Network Leadership and Operations Center (UM1 AI68619), the AIDS Clinical Trials Group Leadership and Operations Center (UM1 AI68636), and the Infectious Diseases Clinical Research Consortium leadership group (UM1AI148684).

Samples from 2019nCoV-307 (ClinicalTrials.gov: NCT05463068) were included in the present study. 2019nCoV-307 was funded by Novavax, Inc. with support from the US BARDA (Contract W15QKN-16-9-1002, Project Number MCDC2011-001).

## References

1. Rispens T, Huijbers MG. The unique properties of IgG4 and its roles in health and disease. Nat Rev Immunol 2023;23(11):763–778. DOI: 10.1038/s41577-023-00871-z.

2. Vidarsson G, Dekkers G, Rispens T. IgG subclasses and allotypes: from structure to effector functions. Front Immunol 2014;5:520. DOI: 10.3389/fimmu.2014.00520.

3. Kober C, Manni S, Wolff S, et al. IgG3 and IgM Identified as Key to SARS-CoV-2 Neutralization in Convalescent Plasma Pools. PLoS One 2022;17(1):e0262162. DOI: 10.1371/journal.pone.0262162.

4. Irrgang P, Gerling J, Kocher K, et al. Class switch toward noninflammatory, spike-specific IgG4 antibodies after repeated SARS-CoV-2 mRNA vaccination. Sci Immunol 2023;8(79):eade2798. DOI: 10.1126/sciimmunol.ade2798.

5. Yoshimura M, Sakamoto A, Ozuru R, et al. The appearance of anti-spike receptor binding domain immunoglobulin G4 responses after repetitive immunization with messenger RNA-based COVID-19 vaccines. Int J Infect Dis 2023;139:1–5. DOI: 10.1016/j.ijid.2023.11.028.

6. Routhu NK, Stampfer SD, Lai L, et al. Efficacy of mRNA-1273 and Novavax ancestral or BA.1 spike booster vaccines against SARS-CoV-2 BA.5 infection in non-human primates. Sci Immunol 2023:eadg7015. DOI: 10.1126/sciimmunol.adg7015.

7. Buhre JS, Pongracz T, Kunsting I, et al. mRNA vaccines against SARS-CoV-2 induce comparably low long-term IgG Fc galactosylation and sialylation levels but increasing long-term IgG4 responses compared to an adenovirus-based vaccine. Front Immunol 2022;13:1020844. DOI: 10.3389/fimmu.2022.1020844.

8. Uversky VN, Redwan EM, Makis W, Rubio-Casillas A. IgG4 Antibodies Induced by Repeated Vaccination May Generate Immune Tolerance to the SARS-CoV-2 Spike Protein. Vaccines (Basel) 2023;11(5). DOI: 10.3390/vaccines11050991.

9. Selva KJ, van de Sandt CE, Lemke MM, et al. Systems serology detects functionally distinct coronavirus antibody features in children and elderly. Nat Commun 2021;12(1):2037. DOI: 10.1038/s41467-021-22236-7.

10. Goldblat D, Alter G, Croty S, Plotkin SA. Correlates of protection against SARS-CoV-2 infection and COVID-19 disease. Immunol Rev 2022;310(1):6–26. DOI: 10.1111/imr.13091.

